# Genome-wide epistasis analysis in Parkinson’s disease between populations with different genetic ancestry reveals significant variant-variant interactions

**DOI:** 10.1101/2022.07.29.22278162

**Authors:** Alejandro Cisterna-Garcia, Bernabe I. Bustos, Sara Bandres-Ciga, Thiago P. Leal, Elif I. Sarihan, Christie Jok, Cornelis Blauwendraat, Mike A. Nalls, Dimitri Krainc, Andrew B. Singleton, International Parkinson’s Disease Genomics Consortium (IPDGC), Ignacio F. Mata, Steven J. Lubbe, Juan A. Botia

## Abstract

Genome-wide association studies (GWAS) have increased our understanding of Parkinson’s disease (PD) genetics through the identification of common disease-associated variants. However, much of the heritability remains unaccounted for and we hypothesized that this could be partly explained by epistasis. Here, we developed a genome-wide non-exhaustive epistasis screening pipeline called *Variant-variant interaction through variable thresholds* (*VARI3*) and applied it to diverse PD GWAS cohorts. First, as a discovery cohort, we used 14 cohorts of European ancestry (14,671 cases and 17,667 controls) to identify candidate variant-variant interactions. Next, we replicated significant results in a cohort with a predominately Latino genetic ancestry (807 cases and 690 controls). We identified 14 significant epistatic signals in the discovery stage, with genes showing enrichment in PD-relevant ontologies and pathways. Next, we successfully replicated two of the 14 interactions, where the signals were located nearby *SNCA* and within *MAPT* and *WNT3*. Finally, we determined that the epistatic effect on PD of those variants was similar between populations. In brief, we identified several epistatic signals associated with PD and replicated associations despite differences in the genetic ancestry between cohorts. We also observed their biological relevance and effect on the phenotype using *in silico* analysis.

## Introduction

Parkinson’s disease (PD) is a complex neurodegenerative disorder stemming from the interaction between genetic and environmental factors ^1^. Some monogenic causes of PD have been identified in familial and early-onset patients, although these account for a small number of cases ^2,3^. The underlying cause of the majority of PD cases, called sporadic PD, is currently unknown. However, genome-wide association studies (GWAS) have done a crucial effort in helping us to understand the etiology of PD identifying 90 disease-associated common variants to date in European populations and 2 additional variants in Asian population, giving light to the biological mechanisms and heritable components of the disease ^4–7^. Nevertheless, GWAS identified variants only explain a small proportion, about 1/3, of the total genetic factors associated with the disease ^8^. There are other possible factors accounting for this missing heritability such as rare variants, structural variants, and genetic interactions ^9^, which are known to be challenging to study, both requiring large sample sizes and specialized equipment and methods to call/analyze them. Here, we aim to assess the effect of interactions (or epistasis) between variants and their association with PD risk ^10–12^.

Studies have suggested that epistasis may play a significant role in neurodegenerative diseases such as Alzheimer’s Disease (AD) ^13,14^, therefore we hypothesized that epistasis might also play a role in PD. Previous PD epistasis studies have generally only assessed interactions between known important variants/genes under specific hypotheses ^15,16^. Shi *et al*. 2016 identified potential epistatic interactions between *GBA* and *LRRK2* that were associated with PD risk in a Chinese cohort ^15^, however, they failed to replicate these findings in an independent cohort. Additionally, Fernandez-Santiago *et al*. 2019 identified potential epistatic interactions between *SNCA-RPTOR-RPS6KA2* associated with reduced age at onset (AAO) of PD ^16^.

In this study, we focused on statistical epistasis, *i*.*e*., the departure from additive effects of genetic variants with regard to their global contribution to the phenotype measured in the population ^17,18^. We limited our assessment to epistatic interactions that involve two single nucleotide polymorphisms (SNPs) that show a statistically significant relevant effect on the phenotype when appearing together in a large-scale and unbiased manner ^19,20^. Studies assessing epistasis within model organisms and artificial cohorts have demonstrated that the most prominent epistatic effects detected have involved variants with high and similar minor allele frequency (MAF > 0.05) ^21^. We have therefore developed a hypothesis-free pipeline, called *Variant-variant interaction through variable thresholds* (*VARI3*), to facilitate the interrogation and identification of variant-variant interactions at a genome-wide level through the inclusion of high MAF SNPs. Here we have successfully applied it to several existing PD GWAS data across diverse populations including 14 independent cohorts of European descent from the International Parkinson’s Disease Genomics Consortium (IPDGC) ^5^ and the Latin American Research Consortium on the Genetics of PD (LARGE-PD) study ^22^ with individuals of Latino ancestry. To facilitate epistatic analysis for the general public, *VARI3* is an available R package that automates the selection and analysis of the most promising SNPs to identify and interpret the results (https://github.com/alexcis95/VARI3).

## Materials and methods

### GWAS cohorts used in the study (Genotyping and Quality-Control Analyses)

We organized our epistasis screen into two stages: using a combined dataset consisting of 14 IPDGC cohorts as a discovery stage to identify candidate statistically significant variant-variant interactions and the LARGE-PD cohort as a replication stage (**Figure 1**).

**Figure 1.**
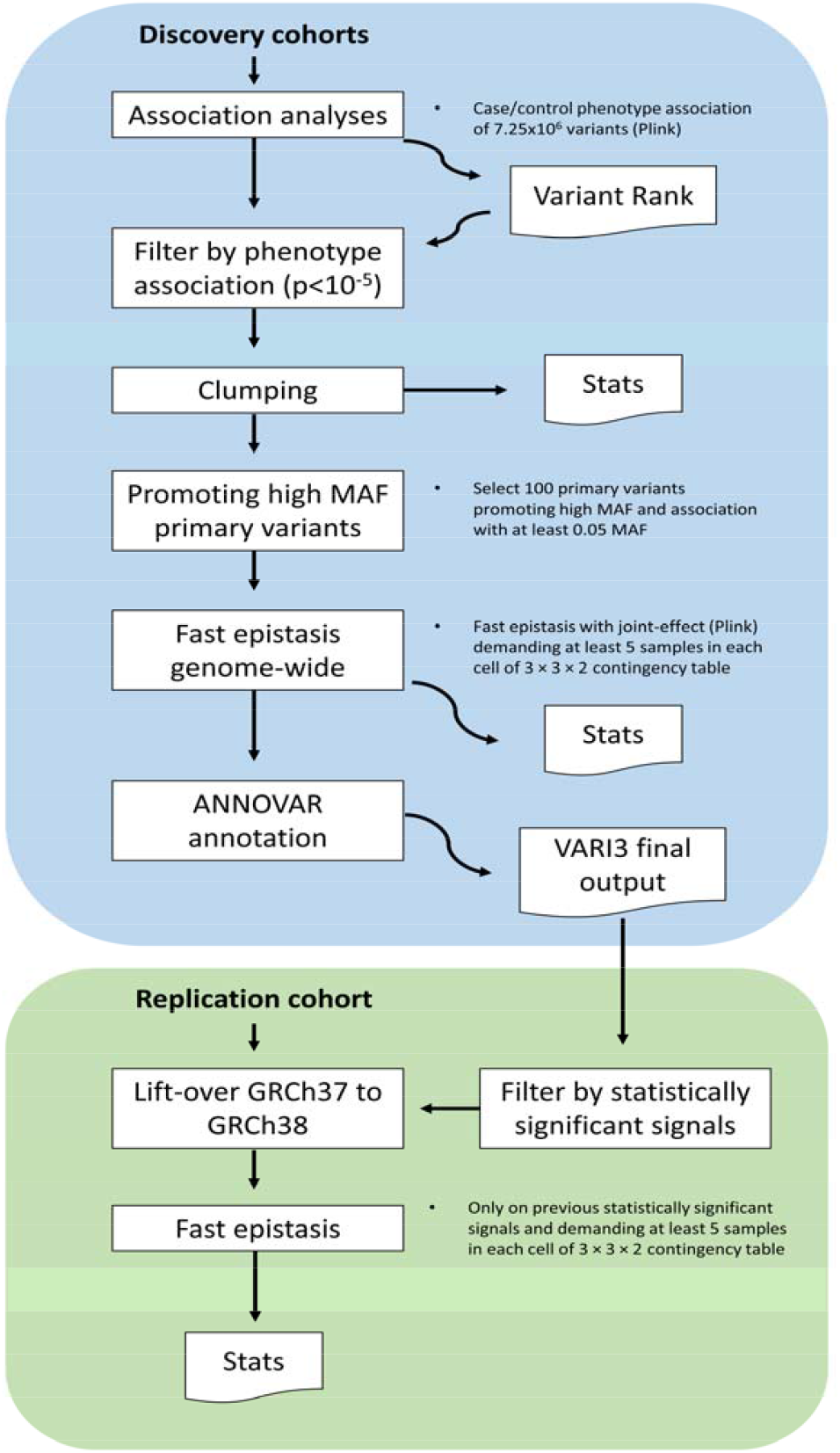
*VARI3* pipeline and study design summary. The blue rectangle shows the *VARI3* pipeline from discovery (IPDGC GWAS) data to final output. The green rectangle shows the steps to evaluate the statistically significant epistatic signals detected in discovery cohorts in the replication cohort.

#### IPDGC GWAS cohorts

The IPDGC-GWAS data consisted of 14 independent cohorts, including 14,671 PD cases and 17,667 healthy controls. All individuals provided informed consent for participation in genetics studies, which were approved by the relevant local ethics committee for each cohort used. The cohorts used and their respective sample sizes are shown in **Supplementary Table 1**. We did not include cohorts genotyped on the NeuroX array to ensure good variant coverage across cohorts (>85%). Briefly, for sample QC prior to imputation, individuals with low call rate, discordance between genetic and reported sex, family relatedness (pi_hat score > 0.125), heterozygosity outliers and ancestry outliers were removed. For genotype QC, variants with a missingness rate of >5%, MAF□<0.01, exhibiting deviations from Hardy–Weinberg Equilibrium (HWE)□<1×10^−5^ and palindromic SNPs were excluded. Hard call genotypes were obtained using plink default value of 0.8 to the variant dosage. Quality control (QC) and genotype imputation were previously described^5^.

#### LARGE-PD cohort

A total of 1,497 individuals (807 PD cases and 690 controls) were recruited from Uruguay, Peru, Chile, Brazil, and Colombia from 2007 to 2015 as part of the LARGE-PD study^22^. The cohorts used and their respective sample sizes are shown in **Supplementary Table 2**. All participants provided written informed consent according to their respective national requirements. Genotyping was performed using the Multi-Ethnic Genotyping Array (MEGA; Illumina). For genotype QC, variants with a missingness rate of >5%, exhibiting deviations from Hardy–Weinberg Equilibrium (HWE)□<1×10^−6^ in controls and <1×10^−10^ in cases. Unaligned, i.e., marks as chromosome 0, duplicated, non-autosomal, and monomorphic SNPs were excluded before filtering. More clinical details and QC procedures have previously been described ^22^.

### Epistasis pipeline and risk interpretation method

The *VARI3* pipeline automates the selection and analysis of the most promising SNPs to identify epistasis and consists of two steps (**Figure 1**). In the first step, we generate a set of primary SNPs to include in the binary epistatic association tests. Here, we performed an association analysis of all SNPs against the phenotype under an additive model using Plink (v1.90b4.4) ^23^, this analysis in our case included as covariates age at onset, sex, and the principal components (PCs) 1-10. To promote the assessment of epistasis, we prioritize all variants with a P <10^−5^ (rather than the accepted genome-wide threshold of P <10^−8^ for the inclusion of variants with higher MAF). This allows us to select SNPs with a superior MAF through the prioritization of high MAF variants (MAF >0.05). We next apply Plink-based clumping to obtain a set of primary SNPs for the top 100 associated loci. We used a default LD window of 250kb and an r^2^ of 0.5 from the index SNP for each locus. In the second step, using the fast epistasis with joint-effect adjusted test in Plink ^24^, which is based on the inspection of 3×3 joint genotype count tables, we tested the set of primary variants against all variants on a genome-wide basis to search for epistatic associations. We used default settings for the fast epistasis with joint-effect run. The p-value for the inclusion of variant pairs in the main report was restricted to 0.0001. Additionally, we used the default quality controls which exclude interactions observed in fewer than five samples, i.e the number of individuals with that particular interaction in 3×3 contingency tables. Then we annotated variants with ANNOVAR ^25^ to obtain gene symbols and genomic localization. Finally, using chi-square statistics and p-values adjusted by the number of tests we obtained the statistically significant variant-variant interactions. The final output is a table with the most statistically significant epistatic interaction for each primary variant pair that has at least a p-value < 10^−5^. Moreover, the *VARI3* package includes the function *TLTO* and therefore automates the conversion of the two locus ratios from Plink to a graph and a table with the odds ratios (ORs) to better understand the epistatic effects in disease. The *two locus* option in Plink generates a file that contains counts and frequencies of the two locus genotypes by cases and controls. *TLTO* uses this file to compute the ORs based on the genotypic OR ^26^, the odds of phenotype (the probability that the phenotype is present compared with the probability that it is absent) in exposed (in a particular genotype combination) *vs*. non-exposed individuals. Finally, *TLTO* generates a graph and a table that shows the phenotypic effects of each genotype combination, *i*.*e*., the ORs with 95% confidence intervals (95% CI) for each genotype combination. The R package is available at https://github.com/alexcis95/VARI3.

### Functional enrichment analysis methods

To functionally characterize the top associated interactions, we carried out loci connectivity analyses across gene-expression datasets from GTEX v.8 ^27^, gene-ontologies and molecular pathways using FUMA ^28^, phenotype enrichment using PhenoExam ^29^, functional gene interaction networks using STRING ^30^, and gene co-expression network analysis using CoExp Web ^31^. SNP lists from significant interactions were extracted for significant eQTL associations in PD-relevant tissues (caudate, putamen, nucleus accumbens and substantia nigra) in the GTEX v.8 data to obtain significant eQTL genes (eGenes). Significant eGenes were analyzed for functional enrichment using FUMA gene2func tool for gene-ontology and pathway enrichments from the Molecular Signature Database v.7.0 ^32^; and functional protein interactions using STRING with median confidence networks (confidence score >400), multiple testing correction was done using the Benjamini-Hochberg FDR method ^33^. Significant eGenes were also analyzed for phenotype enrichment using PhenoExam with Human Phenotype Ontology (HPO) terms ^34^; and gene co-expression network analysis using CoExp Web with GTEX v.7 and substantia nigra tissues, multiple testing correction was done using the Bonferroni method ^35^.

## Results

### Discovery stage in IPDGC GWAS cohorts

From the combined set of genotypes from the 14 PD GWAS cohorts, we were left with 7,258,166 variants and 32,338 samples (14,671 PD cases and 17,667 controls) after individual and variant level quality controls. Using default settings (**Figure 1**), we ran *VARI3* and detected a total of 95 variant-variant interactions, of which 69 were interchromosomal and 26 were intrachromosomal (**Figure 2, Supplementary Table 3**). After multiple testing corrections (7.47×10^6^ valid tests), we observed 14 statistically significant intrachromosomal local (less than 1 mb apart) variant-variant interactions (**Table 1**), some of which include variants that are nearby or within known PD genes and/or GWAS loci such as *SNCA* and *MAPT*. 85.71% (12/14) of the observed interactions involved variants that individually confer very small increased risk or have no effect on PD risk, but when inherited together, result in significantly larger effects on PD risk with 42.86% (6/14) increasing PD risk more than 3-fold. The most prominent of which can be seen for the SNPs located at 15:58980985 (*ADAM10*) and 15:58856033 (*LIPC*) - individually these SNPs had ORs close to 1 (1.14 and 0.99 respectively) but together confer a substantial 7.42-fold [95% CI = 3.13, 17.58] increase in risk.

**Figure 2.**
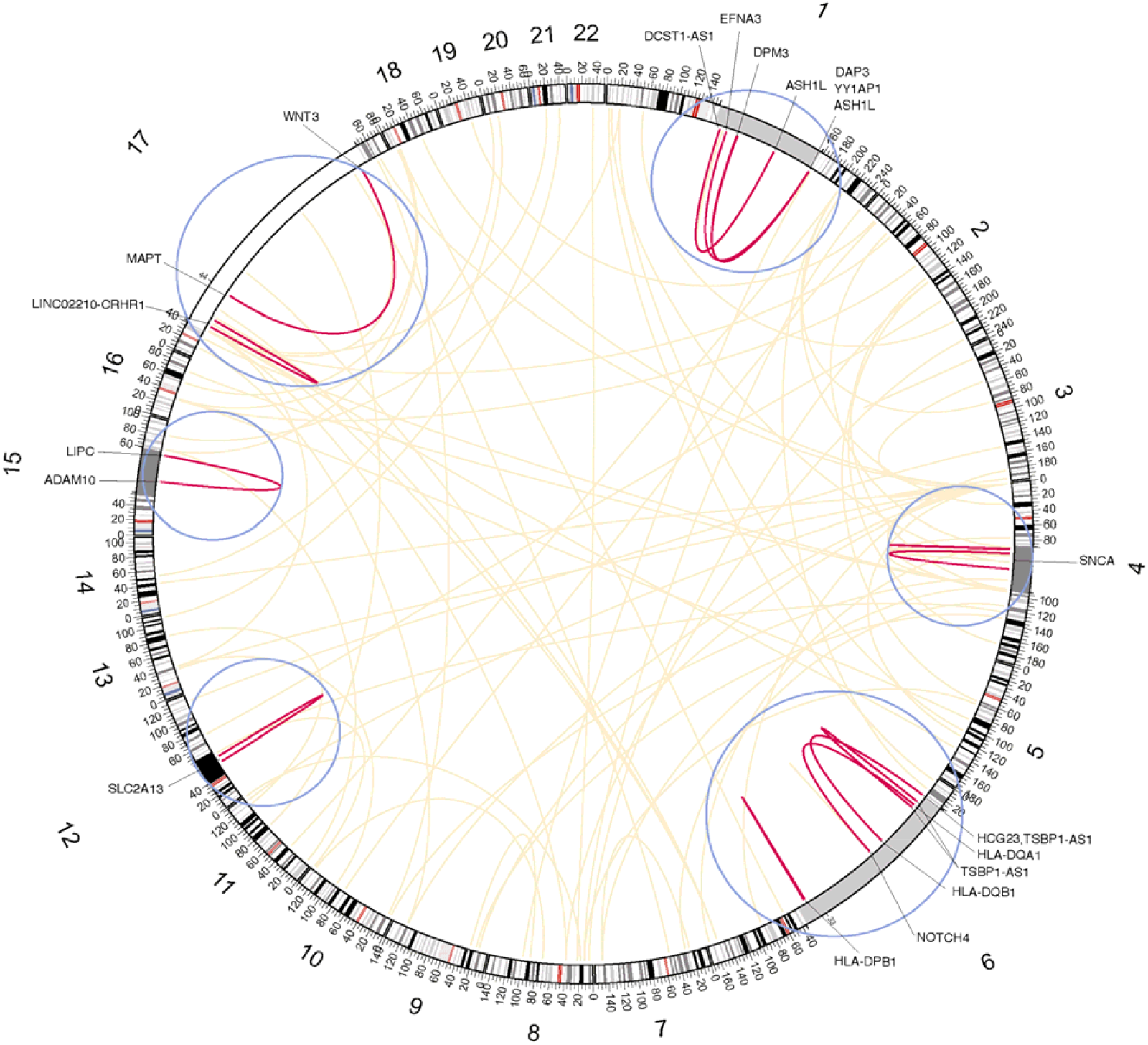
Genome wide distribution of SNP-SNP interactions observed across IPDGC GWAS discovery cohorts. Circos plot showing the 95 interactions obtained from *VARI3* across chromosomes 1-22 from the IPDGC GWAS cohorts. Yellow links show non-significant interactions. Blue circles show 300x zoomed-in regions, containing red links depicting the 14 significant interactions after Bonferroni correction and their nearby gen

**Table 1.**
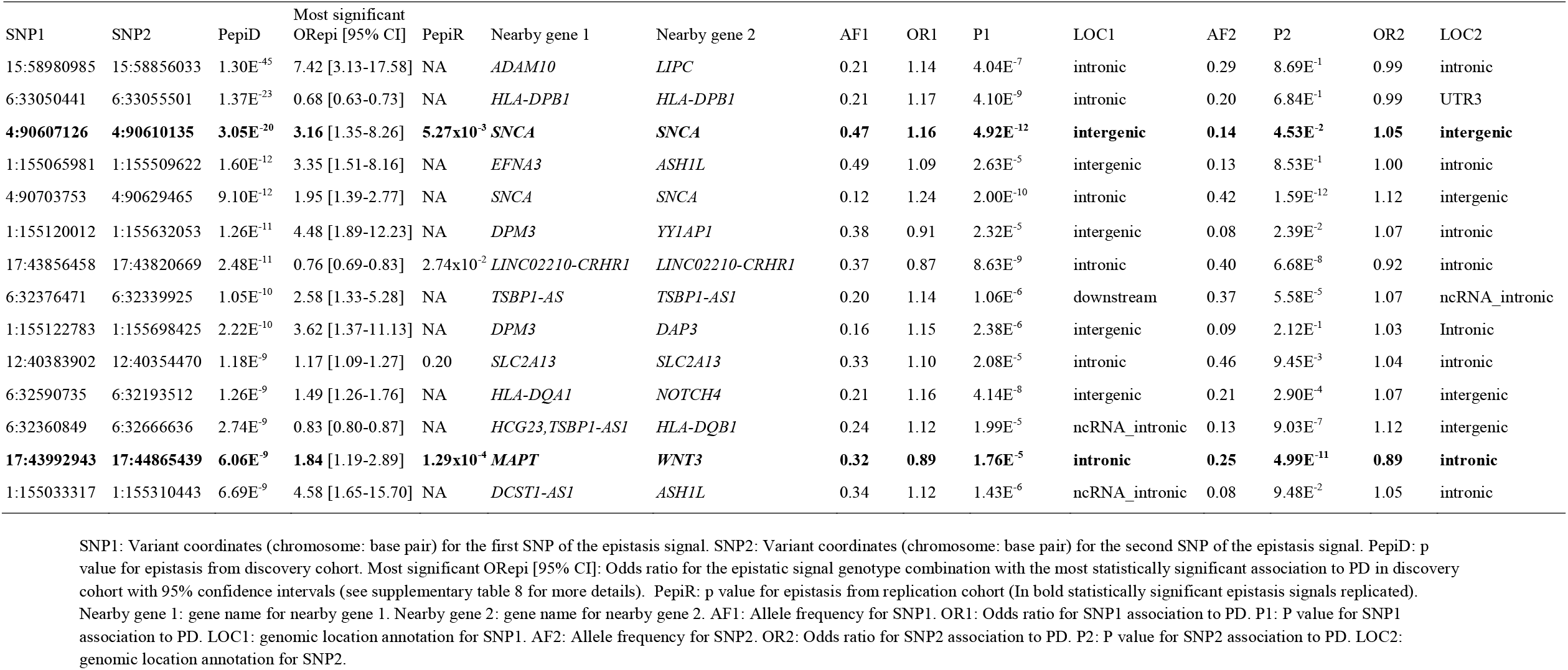
Statistically significant interactions from the discovery and replication cohorts.

### Effect of interacting SNPs on gene expression and functional enrichment analyses of the significant interactions

To explore functional insights from the 14 genome-wide significant interactions found in the discovery stage, we extracted moderate to high LD variants (r^2^ >0.5) in 1Mb from each of the 28 interacting SNPs, obtaining 134 variants for further analyses (**Supplementary Table 4**). Using gene eQTL information from GTEx v.8 we found 84 of the 134 variants have significant eQTLs in PD-relevant tissues: 51 in the caudate (367 variant-gene pairs), 60 in putamen (322 variant-gene pairs), 58 in the nucleus accumbens (362 variant-gene pairs), and 37 in the substantia nigra (225 variant-gene pairs) (**Supplementary Table 5**). The identified significant eQTLs were seen to target a total of 30 unique nearby genes: 26 in the caudate, 19 in putamen, 23 in the nucleus accumbens, and 12 in the substantia nigra, including genes within known PD GWAS loci (*KANSL1, CRHR1* and *MAPT*) and *HLA* genes (*HLA-DOA, HLA-DRA, HLA-DPB1, HLA-DQB2* and *HLA-DQA2*).

We next used the 30 eQTL-nominated genes and performed gene ontology, pathway, phenotype, and network enrichment analyses to further dissect the interactions’ functional connections. Gene ontology analyses showed significant enrichments in categories related to the immune system, mostly driven by the *HLA* genes shown above, and include antigen processing and presentation of peptide antigen via MHC class II (GO:0002495), T-cell receptor signaling pathway (GO:0050852), and Interferon Gamma signaling pathway (GO:0060333) (**Supplementary Figure 1**). Similarly, the pathway enrichment analysis showed significant enrichment in molecular pathways related to the immune system: Asthma (KEGG H00079), Intestinal immune network for IGA production (KEGG HSA04672), and Translocation of ZAP-70 to Immunological synapse (Reactome HSA202430) (**Supplementary Figure 2**). The phenotype enrichment analysis in PhenoExam showed significant enrichment in HPO terms associated with PD: Weight loss (HP:0001824), Diplopia (HP:0000651), and Substantia nigra gliosis (HP:0011960) (**Supplementary Figure 3 and Supplementary Table 6**). Additionally, the gene co-expression network analysis using CoExp Web with GTEX v.7 and substantia nigra tissue showed significant eGene overlap in the green module (p =0.02) (**Supplementary Table 7**). We found the following eGenes in the green module: *SNCA, SLC2A13* and *GPRIN3*; the green module is associated with dopaminergic neurons and dopamine transport (p=8.92×10^−3^). Network enrichment analysis in STRING showed significant connections (p <1×10^−6^) of the *HLA* genes and the *MAPT* locus, including *KANSL1, LRRC37A2, MAPT* and *CRHR1* (**Supplementary Figure 4**). Overall, these results indicate that the significant interacting SNPs influence gene expression and are enriched in PD-relevant ontologies, phenotypes, and pathways.

### Replication in LARGE-PD cohort

We tested the 14 significant variant-variant interactions identified in the IPDGC cohorts using the LARGE-PD. First, we lifted over the 14 interactions from GRCh37 to GRCh38 using the Lift Genome Annotations tool ^36^ to match the assembly version in the replication cohort. Only four of the 14 interacting variants from the discovery stage survived our quality controls in the replication dataset, where interactions involving fewer than 5 samples in one of the nine possible genotype variant-variant combinations were excluded. Thus, the Bonferroni-adjusted threshold in the replication cohort is 1.25×10^−2^ (0.05/4). We could only replicate two of the statistically significant variant-variant interactions (bold letters in **Table 1**) - the *SNCA-SNCA* (4:90607126 - 4:90610135; p=5.27×10^−3^) and the *MAPT-WNT3* interaction (17:43992943 - 17:44865439; p=1.29×10^−4^). The *LINC02210-CRHR1* (p=2.74×10^−2^) and *SLC2A13-SLC2A13* (p=0.20) interactions did not surpass multiple testing correction.

### Understanding the genotype-specific effect in the variant-variant interactions

We used the two-locus ratio analyses to understand the effect on the phenotype of each genotype combination observed in the 14 statistically significant variant-variant interactions that we identified in the IPDGC cohorts (discovery). To facilitate the interpretation, the OR and the 95% CIs for each variant-variant interaction were computed using the *TLTO* function (**Supplementary Table 8**). We observed that the epistatic signal located within *ADAM10* and *LIPC* (15:58980985 - 15:58856033) was the highest risk variant-variant interaction associated with PD (C/C-T/T genotype combination; OR [95% CI]=7.42 [3.13, 17.58]) whereas the highest protective epistatic signal was located within *HLA-DPB1* (6:33050441 - 6:33055501) (G/G-T/C genotype combination; OR [95% CI]=0.68 [0.63, 0.73]). Interestingly, the SNPs at 17:43992943 (*MAPT*) and 17:44865439 (*WNT3*) individually appear to confer reduced risk (OR=0.89 and 0.89 respectively **Table 1**) but through epistasis, the effect of the variant-variant interaction on PD depends on genotype combinations, different genotype combinations resulted in significantly increased PD risk (G/G-G/G combination, OR [95% CI]=1.84 [1.18, 2.89]) or reduced PD risk (A/G-G/T combination, OR [95% CI]=0.85 [0.81, 0.90]).

Focusing on the two statistically significant variant-variant interactions that were replicated in the LARGE-PD cohort (**Figure 3**), we observed similar combined genotype effects for the majority of each genotype combination from the epistatic signals between stages. Interestingly, at the *SNCA-SNCA* locus (4:90607126 - 4:90610135), the G/G-T/T genotype combination was associated with a higher risk for both the discovery and replication cohorts (OR [95% CI]=3.16 [1.35, 8.26] and OR [95% CI]=2.43 [1.61, 3.73] respectively) while the G/G-A/A genotype combination was associated with reduced risk in carriers (OR [95% CI]=0.77 [0.74, 0.81] and OR [95% CI]=0.71 [0.56, 0.91] respectively). For the *MAPT* and *WNT3* interaction (17:43992943 - 17:44865439), the G/G-G/G combination was associated with higher risk in both discovery and replication cohorts (OR [95% CI]=1.84 [1.19, 2.89] and OR [95% CI]=1.74 [1.24, 2.46] respectively) while the A/G-G/T genotype combination was associated with reduced risk in carriers (OR [95% CI]=0.85 [0.81, 0.90] and OR [95% CI]=0.71 [0.54, 0.95] respectively).

**Figure 3.**
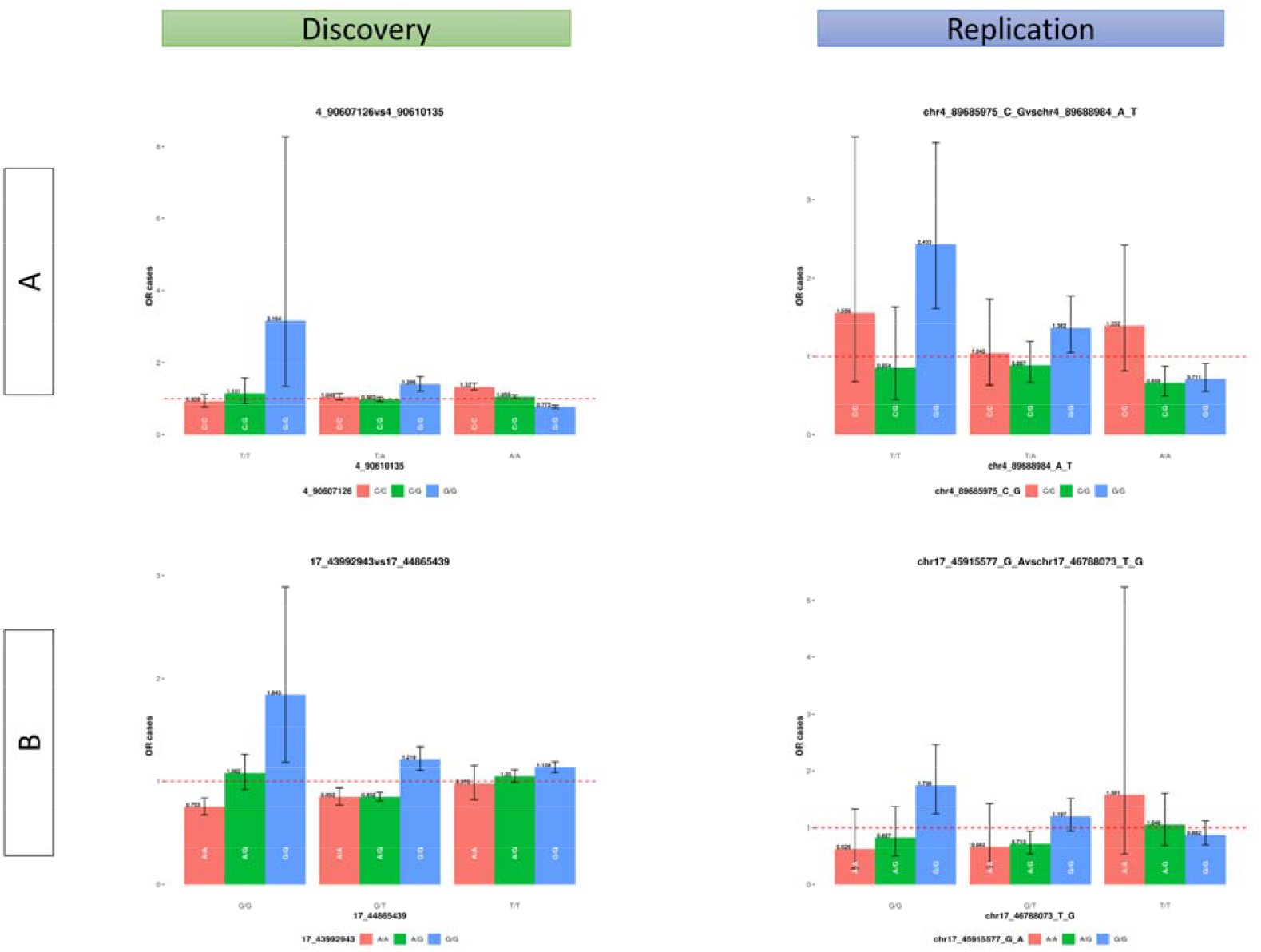
Odds ratio of replicated variant-variant interactions of the two locus genotypes. A) Replicated interaction located in chromosome 4 by intergenic variants nearby *SNCA* (Genome Reference GRCh37, 4:90607126 - 4:90610135, Pint=5.27×10^−3^). B) Replicated interaction located in chromosome 17 within *MAPT* and *WNT3* (Genome Reference GRCh37, 17:43992943 - 17:44865439, Pint=1.29×10^−4^). Each bar indicates the odds ratio (OR) in the y-axis for each genotype combination from the epistatic signals. Each colour indicates the genotype from one SNP and the x-axis indicates the genotype from the other SNP in the epistatic signals. The horizontal dash line indicates the OR is equal to 1. The black vertical line defines the 95% confidence interval for each genotype combination from the epistatic signals.

## Discussion

Here we present the newly developed *VARI3* pipeline that performs non-exhaustive genome-wide epistasis screens. To demonstrate its utility, we successfully applied this pipeline to 14 independent PD datasets from the IPDGC and identified 14 significant epistatic signals (**Table 1**). Next, we evaluated them in the replication stage using a Latino ancestry cohort finding two significant interactions in the *SNCA* locus and within the *MAPT* and *WNT3* loci. Functional *in silico* findings revealed significant enrichment of pathways related to the immune system, phenotypes and coexpression modules associated with PD (**Supplementary Figure 1-4** and **Supplementary Table 4-7**). Finally, we determined that the epistatic effect on PD of those variants was similar between populations by showing that risk profiles associated with different genotype combinations followed similar patterns (**Figure 3** and **Supplementary Table 8**).

Epistasis studies at a genome-wide level have been a challenge due to exhaustive pairwise testing of millions of variants and reduced genetic power due to the lack of large sample sizes. Typically, epistasis studies in human complex neurodegenerative diseases are hypothesis-driven ^13,15,16^ to reduce the multiple-testing correction burden resulting from the large number of tests conducted in genome-wide scans ^14^. The number of multiple tests can be reduced using different approaches such as looking at genes of interest based on previous biological knowledge in AD and PD ^13,15^. This approach is simplistic and reduces the epistasis complexity by limiting the screen to a few SNPs. Another method is to restrict SNP selection to those within relevant biological/functional pathways ^16^. Finally, Wang *et al*. 2021 limited their genome-wide epistasis scan to SNPs that were predicted to be probably damaging (CADD scores ≥15) ^14^. Although this is not an exhaustive search for epistasis interactions, they selected 36,860 SNPs and performed ∼3.92×10^8^ valid tests. Similarly, *VARI3* reduces the number of tests to achieve enhanced statistical power in our genome-wide epistasis scan by prioritizing SNPs based on high MAF, low LD – to ensure the inclusion of independent SNPs – and all variants with a P <10^−5^ associated with the disease, selecting 100 primary variants for further analysis. This allowed us to identify new local epistasis between variants within known PD loci including *SNCA, MAPT* and *CRHR1* ^15,16^. Our results demonstrate that using variant prioritization methods increases the power to detect novel genome-wide significant variant-variant interactions in disease-relevant genes.

Functional insights from the 14 interactions obtained in the discovered stage and their high-LD neighboring variants revealed significant enrichment in pathways related to the immune system. This finding is in line with Bandres-Ciga *et al*. 2020 ^37^ who highlighted the immune system response as one of the main contributors to PD etiology. From the gene expression analyses and the ontology and pathway enrichment analysis, we observe significant results driven by the *HLA* locus, with four variants among the significant interactions observed in the discovery stage. Due to the highly complex haplotype structure of the *HLA* locus, being the most gene dense and most polymorphic part of the human genome, multiple studies remove this region from the analysis ^38–40^. However, previous epistatic studies have analyzed this region finding significant interactions, determining genetic susceptibility in multiple sclerosis ^41^, and Takayasu arteritis ^42^. The involvement of the *HLA* locus in the immune system and their association with PD is well documented ^43–46^. Here we found epistatic interactions between the *HLA* genes *HLA-DPB1, HLA-DQA1*, and *HLA-DQB1* which have been found to have SNPs with genome-wide significant associations in the latest PD GWAS meta-analysis ^5^, Adding further evidence to the role that *HLA* genes and the immune pathways play in PD etiology.

Replication in an independent cohort is highly recommended in any GWAS study, but especially in variant-variant interaction studies due to false-positive results stemming from the great magnitude of hypotheses being tested, and false negatives caused by poor statistical power ^47^. The majority of epistasis studies do not cover the replication issue which is essential to obtaining reliable results ^48^. To overcome the above, we prioritized high MAF variants across the discovery and replication cohorts consisting of 32,338 and 1,497 individuals respectively, in order to ensure a high overlap of SNPs between datasets. Using this approach, we replicated two variant-variant interactions: one in the *SNCA* locus (4:90607126 - 4:90610135) and the other within *MAPT* and *WNT3* (17:43992943 - 17:44865439). *SNCA, MAPT* and *WNT3* are well-known PD risk genes, with multiple SNPs identified as risk factors for the disease ^49–52^. Besides highlighting the interactions, the risk interpretation of the variant-variant genotype combinations is key in understanding their effect size on the individuals. To the best of our knowledge, no tool exists that allows an easy interpretation of the effect sizes obtained from different genotype combinations and their impact on the individuals. We here developed *TLTO* to automate this task, where the tool computes the OR for each genotype combination in a 3□×□3□contingency table, using cases and controls to produce a graph for easy assessment of the epistatic associations. Our findings with *TLTO* show that the effect size of individual SNPs is smaller than when both variants epistatically interact, which is consistently observed in both discovery and replication stages despite having different ancestry compositions. This suggests that the European ancestry proportion in the replication cohort could be playing a relevant part in the associations observed and that further admixture analysis is needed in order to support this claim.

Genetic ancestry plays an important role in genetic studies, especially when the genetic architecture differs between samples and results obtained from one population may not generalize to others ^53–56^. The detection of epistatic signals could be affected by the possible existence of complex higher-order (>2 SNPs) interactions, genetic heterogeneity, and varying patterns of genetic architecture ^57^. Therefore, genetic ancestry differences could also affect these factors and mask epistatic signals. We have observed that there are slight differences in the effect sizes magnitudes associated with PD of those variant-variant interactions between cohorts, showing a stronger effect in the discovery stage that is composed of individuals of European ancestry. Recent admixture in Latino populations shows a significant proportion of European ancestry, suggesting that the replication results observed in our study could have been affected by such ancestry proportion.

The sample size required to obtain reliable results in epistasis analyses is large and it is affected by variant allele frequency, epistatic effect strength, population prevalence and study design ^58–60^. Furthermore, it is worth mentioning that statistical epistasis may not be the same as biological epistasis, thus targeted approaches that aim to reduce epistasis screening to a very low number of tests have been more suitable due to the lack of larger sample sizes needed for GWAS scans ^12,47^. Although our epistasis study utilizes one of the largest GWAS datasets in PD and we prioritized 100 primary SNPs to test for interactions, the sample size is still a limitation. It was not possible to test 10 of 14 statistically significant epistatic signals detected in the discovery stage in the replication stage because in the latter we could not obtain enough observations of genotype combinations for the selected SNPs. To assess the sample size required to replicate all 14 statistically significant variant-variant interactions, we focused on the rarest epistatic genotype combination (*DPM3*-*DAP3*, genotype combination G/G-T/T, number of PD cases=18 and controls=6). Assuming similar genotype distribution between cohorts, we determined that 4,075 PD cases and 14,722 controls (19,797 individuals) are needed in the replication cohort. With regard to how these epistatic signals affect PD risk prediction, our results suggest that when inherited together, those variant-variant interactions have larger effects on PD risk. We observed that 42.86% (6/14 statistically significant epistatic signals) increased PD risk more than 3-fold compared to the individual SNPs. Therefore, it could be essential to include them and study their effect on PD polygenic risk scores (PRS), similarly to Wang et al. 2021. However, to establish the new PRS with the epistatic interactions we need another independent dataset with enough sample size. Thus, this needs to be addressed in further studies, in order to understand the contribution of epistatic interactions in PD risk prediction and to help in PD patient risk assessment.

In summary, here we have described several limitations and advantages of using *VARI3* and our two-stage study design. Despite the difference in genetic ancestry, our methods allowed us to identify 14 significant epistatic signals associated with PD and replicated two of them in an independent cohort. We also showed how the significant interactions are enriched in PD-relevant pathways. Finally, we determined the statistical effect on the phenotype of those variants and observed similar effects on the phenotype of those interactions in both stages. Our results show that epistatic interactions are contributing with extra risk or protective effect to PD compared to individual variants, therefore helping to reduce part of the missing heritability in the disease and providing a base for larger genome-wide epistatic studies to uncover more interacting variants and genes.

## Data Availability

The code generated during this study is available at https://github.com/alexcis95/VARI3 Data used in preparation of this article were obtained from controlled access data from the IPDGC, including the Baylor College of Medicine / University of Maryland cohort, Finnish PD GWAS, McGill Parkinson's cohort, Oslo PD Study, Spanish Parkinson's cohort, UK PD GWAS, Vance PD cohort, Dutch PD GWAS, German PD GWAS, NIA PD GWAS, PROPARK study, PROBAND study, TUBI (Tubingen) cohort, Myers-Faroud cohort, and LARGE-PD cohort. As the analyses in this manuscript utilize secondary analyses of suitably anonymized datasets, they do not require ethics committee review. The respective ethical committees for medical research approved involvement in genetic studies and all participants gave written informed consent in the original publications for the datasets used. The details of these studies can be obtained upon contacting the IPDGC.

## Declaration of interest

D.K. is the Founder and Scientific Advisory Board Chair of Lysosomal Therapeutics Inc. and Vanqua Bio. D.K. serves on the scientific advisory boards of The Silverstein Foundation, Intellia Therapeutics, AcureX and Prevail Therapeutics and is a Venture Partner at OrbiMed. M.A.N. is the founder and CEO/Consultant of Data Tecnica International LLC, and serves on the scientific advisory board for Clover Therapeutics and is an advisor to Neuron23 Inc. A.C-G., B.I.B, S.B-C., T.P.L, E.I.S, C.J, C.B, A.B.S, I.F.M, S.J.L and J.A.B declare that they have no competing interests.

## Acknowledgments

A.C-G. was supported by the Science and Technology Agency, Séneca Foundation, Comunidad Autónoma Región de Murcia, Spain through the grant 20762/FPI/18. D.K. is supported by the Simpson Querrey Center for Neurogenetics.

## Author contributions

A.C-G., B.I.B., I.F.M., S.J.L and J.A.B. conceived and wrote this article. A.C-G. and J.A.B. conceptualized and designed the *VARI3* software. A.C-G., B.I.B., S.B-C., T.P.L. and E.I.S. processed all the data. All authors participated in the paper writing up, discussed the project, revised the manuscript, and provided critical feedback.

## Web resources

String: https://string-db.org/

GTEX: https://gtexportal.org/

FUMA: https://fuma.ctglab.nl/

CoExpWeb: https://rytenlab.com/coexp/Run/Annotated

## Code and data availability

The code generated during this study is available at https://github.com/alexcis95/VARI3 All genetic data used in this study is available (upon application) from the following sites: (i) International Parkinson’s Disease Genomics Consortium (https://pdgenetics.org/resources). (ii) The Latin American Research consortium on the GEnetics of Parkinson’s Disease (https://large-pd.org/). The details of the IRB/oversight body that provided approval or exemption for the research described are given below: As analyses utilize secondary analyses of suitably anonymized datasets, they do not require ethics committee review. The respective ethical committees for medical research approved involvement in genetic studies and all participants gave written informed consent in the original publications for the datasets used. All necessary patient/participant consent has been obtained and the appropriate institutional forms have been archived.

## Notes

### Funding Statement

A.C-G. was supported by the Science and Technology Agency, Seneca Foundation, Comunidad Autonoma Region de Murcia, Spain through the grant 20762/FPI/18. D.K. is supported by the Simpson Querrey Center for Neurogenetics.

### Author Declarations

Data used in preparation of this article were obtained from controlled access data from the IPDGC, including the Baylor College of Medicine / University of Maryland cohort, Finnish PD GWAS, McGill Parkinson's cohort, Oslo PD Study, Spanish Parkinson's cohort, UK PD GWAS, Vance PD cohort, Dutch PD GWAS, German PD GWAS, NIA PD GWAS, PROPARK study, PROBAND study, TUBI (Tubingen) cohort, Myers-Faroud cohort, and LARGE-PD cohort. As the analyses in this manuscript utilize secondary analyses of suitably anonymized datasets, they do not require ethics committee review. The respective ethical committees for medical research approved involvement in genetic studies and all participants gave written informed consent in the original publications for the datasets used. The details of these studies can be obtained upon contacting the IPDGC.

