## Supplementary Figure 1 for "Genome-wide epistasis analysis in Parkinson’s disease between populations with different genetic ancestry reveals significant variant-variant interactions"

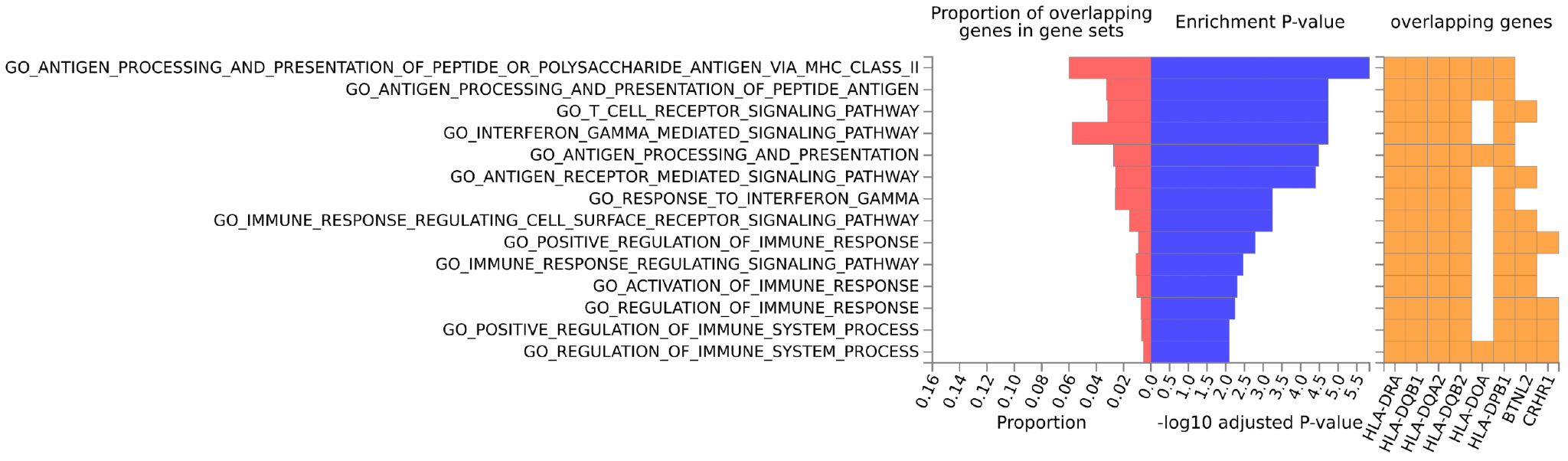

**Supplementary Figure 1. Gene-ontology enrichment of the significant eGenes from interacting variants.** Gene ontology enrichment of genes in FUMA gene2func tool. Pink bars indicate the proportion of overlapping genes with each gene set. Blue bars indicate the -log10 adjusted P value. Yellow squares depict the genes contained in each GO category.
