## Supplementary Figure 2 for "Genome-wide epistasis analysis in Parkinson’s disease between populations with different genetic ancestry reveals significant variant-variant interactions"

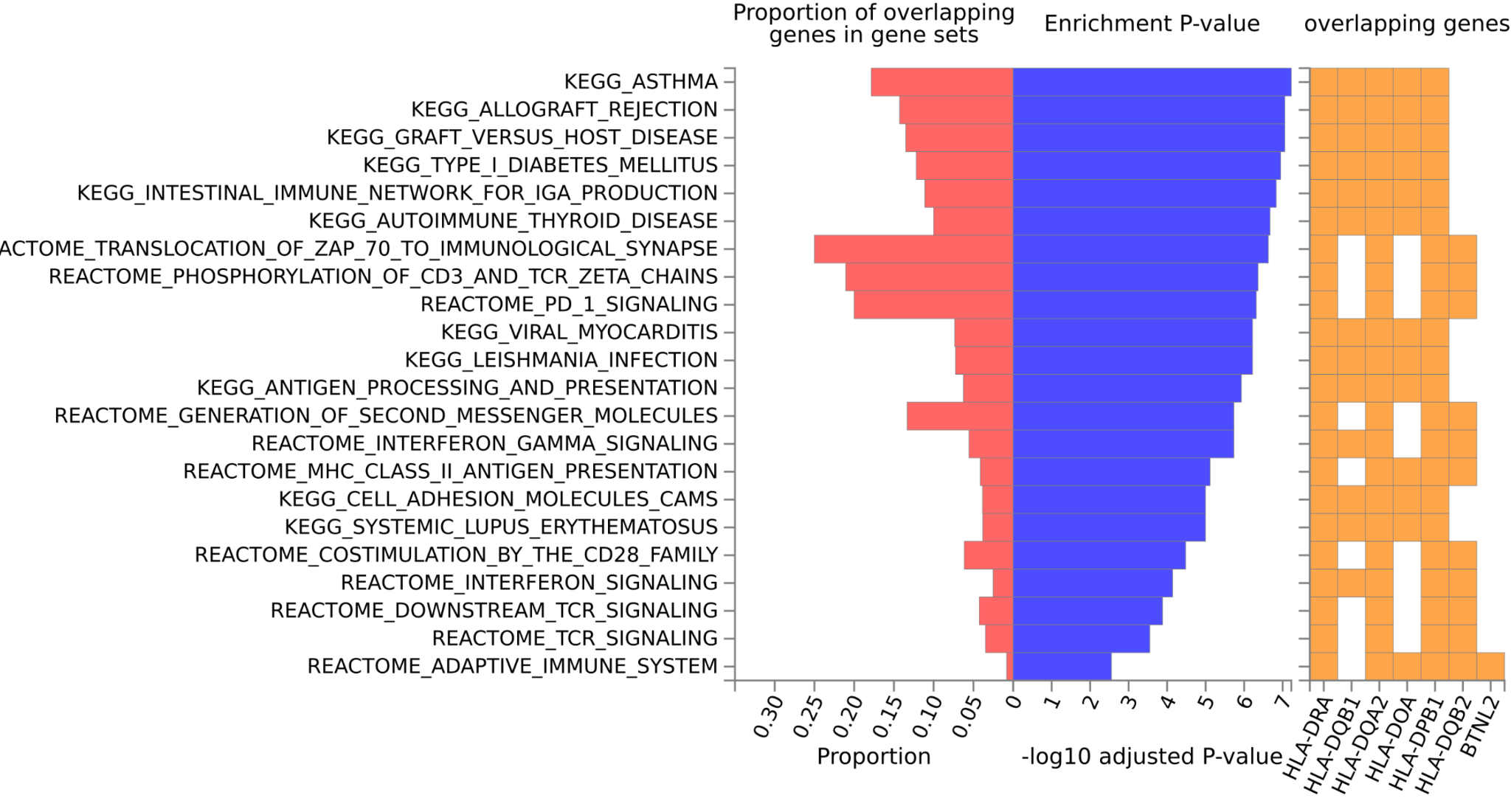
**Supplementary Figure 2. Molecular pathways enrichment of the significant eGenes from interacting variants.** Molecular pathway enrichment of genes in FUMA gene2func tool. Pathway categories were used from the MsigDB database. Pink bars indicate the proportion of overlapping genes with each gene set. Blue bars indicate the -log10 adjusted P value. Yellow squares depict the genes contained in each GO category.
