## Supplementary Figure 3 for "Genome-wide epistasis analysis in Parkinson’s disease between populations with different genetic ancestry reveals significant variant-variant interactions"

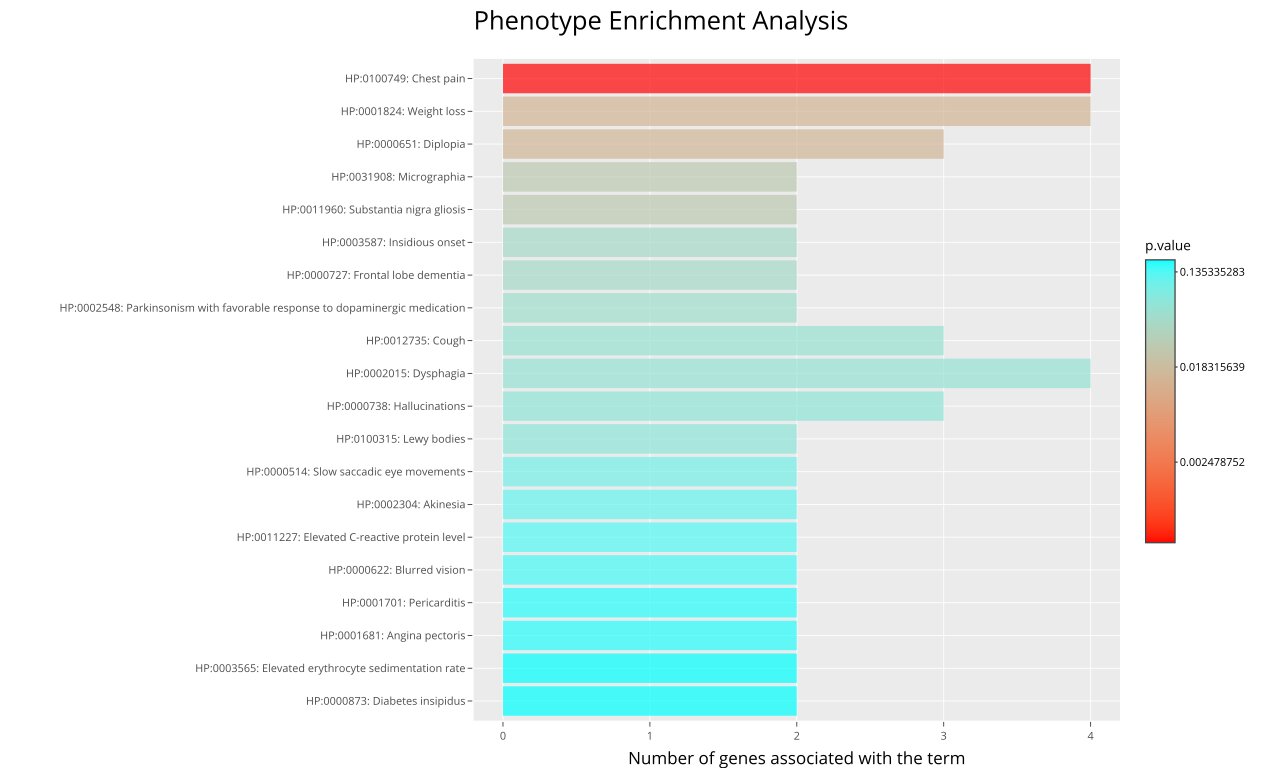

**Supplementary Figure 3. Top 20 significant Human Phenotype Ontology (HPO) terms associated with the eGenes from interacting variants.** Phenotype terms were used from the HPO. X-axis bars indicate the number of eGenes associated with the HPO term and Y-axis indicates the HPO term. Colors of the bars indicate the Bonferroni adjusted P value for each HPO term.
