## Supplementary Figure 4 for "Genome-wide epistasis analysis in Parkinson’s disease between populations with different genetic ancestry reveals significant variant-variant interactions"

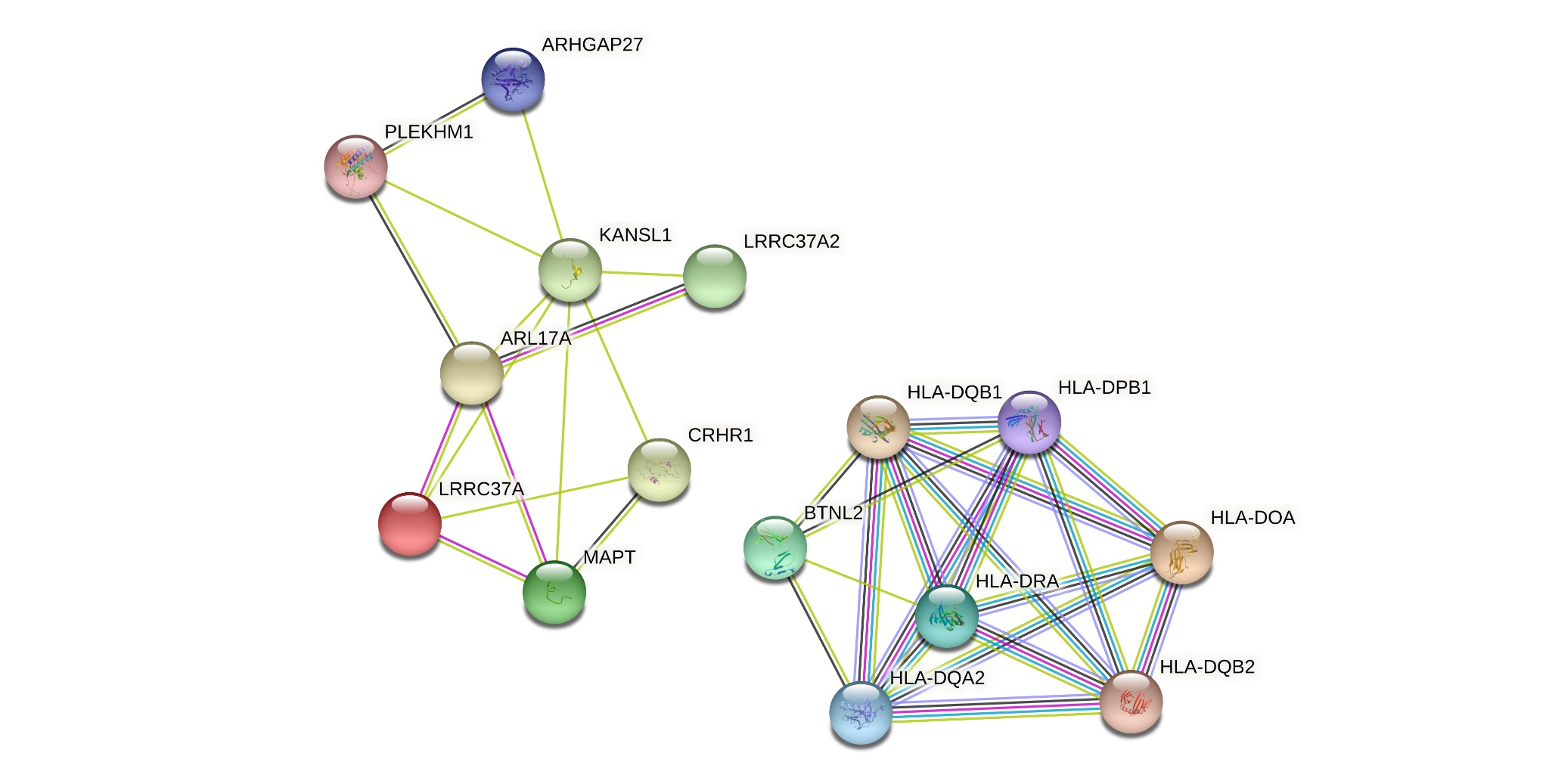

**Supplementary Figure 4. Gene functional interaction networks from eGenes from interacting variants.** The networks were built using STRING using the list of eGenes coming from interacting variants. Each circle represents a gene belonging to a significant network. Links between circles depict the strength of evidence for the gene-gene interaction. P value for network < 1x10^-6^.
